# Cost-effectiveness of endovascular versus open surgery for chronic limb-threatening ischemia

**DOI:** 10.1101/2025.09.22.25336403

**Authors:** Zafar Zafari, Mehdi Najafzadeh, Mufaddal Mahesri, HoJin Shin, Philip P. Goodney, Michael S. Conte, Mark A. Creager, Michael D. Dake, Michael R. Jaff, John A. Kaufman, Richard J. Powell, Chris J. White, Michael B. Strong, Kenneth Rosenfield, Alik Farber, Matthew T. Menard, Niteesh K. Choudhry

**Affiliations:** The University of Maryland-Institute for Health Computing, Bethesda, MD; Department of Practice, Sciences, and Health Outcomes Research at the University of Maryland School of Pharmacy, Baltimore, MD; Medidata Solutions, Boston, MA; Division of Pharmacoepidemiology and Pharmacoeconomics, Department of Medicine, Brigham and Women’s Hospital and Harvard Medical School; Heart and Vascular Center, Dartmouth Hitchcock Medical Center, Geisel School of Medicine at Dartmouth, Lebanon, NH, USA; Division of Vascular and Endovascular Surgery, University of California, San Francisco, San Francisco, CA, USA; Department of Medical Imaging, University of Arizona Health Sciences, Tucson, AZ, USA; Harvard Medical School, Boston, MA, USA (retired); Division of Interventional Radiology, Oregon Health and Science University, Portland, Oregon, USA; Department of Cardiovascular Diseases, The Ochsner Clinical School, New Orleans, LA, USA; Division of Vascular and Endovascular Surgery, Brigham and Women’s Hospital, Harvard Medical School, Boston, MA, USA; Section of Vascular Medicine and Intervention, Massachusetts General Hospital, Harvard Medical School, Boston, MA; Division of Vascular and Endovascular Surgery, Boston Medical Center, Boston University Chobanian & Avedisian School of Medicine, Boston, MA, USA

**Keywords:** Economic evaluation, cost-effectiveness, endovascular surgery, open surgery, limb-threatening ischemia

## Abstract

**Background:** Revascularization for Chronic Limb-Threatening Ischemia (CLTI) may be performed with an endovascular (Endo) or open surgical (Bypass) approach.

**Objective:** To evaluate the cost-effectiveness of Endo versus Bypass surgery for CLTI using data from the Best Endovascular versus Best Surgical Therapy for Patients with CLTI (BEST-CLI) trial.

**Methods:** We developed an individual-level continuous time Markov model that included health states representing the occurrence of adjudicated clinical events from BEST-CLI. Rates of clinical outcomes and health utilities were derived directly from trial data. Costs came from Medicare insurance claims data and physician fee schedule. We calculated the incremental cost per life years gained, incremental quality-adjusted life years (QALYs) gained, incremental net monetary benefit (INMB) and cost per major events of amputation, revascularization, and myocardial infarction (MI) or stroke avoided over a 5– and 10-year time horizon. Sensitivity analyses were performed using a Monte Carlo simulation.

**Results:** In base case analyses conducted over a 5-year time horizon, the mean per person direct medical costs were $227,341 (95% Credible Interval [CrI]: $173,075, $291,443) for Bypass and $243,614 (95% CrI: $190,112, $305,605) for Endo. The mean survival per person was 3.91 years (95% CrI: 3.78, 4.03) for Bypass and 3.88 years (95% CrI: 3.68, 4.06) for Endo. This resulted in Endo being dominated by Bypass surgery with respect to costs per life year gained. The mean QALYs per person were 2.48 (95% CrI: 1.11, 3.49) for Bypass and 2.54 (95% CrI: 1.39, 3.40) for Endo, resulting in an incremental costs per QALY gained of $263,973/QALY and an INMB of –$10,109 (95% CrI: –$168,908, $157,433) at a $100,000/QALY willingness-to-pay threshold for Endo vs. Bypass. The results over 10 years were consistent with those of the 5-year follow-up. In the Monte Carlo simulation, there was only a 55% chance that Bypass was more cost-effective than Endo.

**Conclusion:** In the base case analysis, Bypass was the preferred strategy with respect to survival and QALYs, at conventional willingness to pay thresholds. There was substantial uncertainty around these estimates in probabilistic sensitivity analysis, justifying future research to identify subgroups for whom each of these approaches may definitively be cost-effective.

## BACKGROUND

Chronic Limb-Threatening Ischemia (CLTI) is the most severe manifestation of atherosclerotic lower extremity peripheral artery disease (PAD) (1). Globally, more than 200 million people have PAD (2) with CLTI affecting 11% of these individuals (1). CLTI is associated with a high risk of leg amputation and adverse cardiovascular outcomes, including myocardial infarction, stroke, and cardiovascular death (2,3). CLTI also has substantial economic implications as it results in an estimated annual cost approximating $12 billion in the United States (3).

Along with medical therapy to reduce cardiovascular risk and limb care to control infection and optimize wound healing, treatment of CLTI requires revascularization to improve limb perfusion with either an endovascular (Endo) or open surgical (Bypass) approach (4). The clinical effectiveness of Bypass compared to Endo treatment strategies for CLTI was evaluated in the *Best Endovascular versus Best Surgical Therapy for Patients with CLTI* (BEST-CLI) trial. This study found that among patients who were suitable candidates for surgical bypass with single segment great saphenous vein (SSGSV; designated as “Cohort 1” in the trial) those randomized to Bypass had a 33% risk reduction of the primary outcome, the composite of all-cause death or major adverse limb events (MALE), defined as above-ankle amputation or major limb reintervention, compared to those in the Endo arm, as well as significant reductions in the secondary endpoints of above-ankle amputation and major reintervention (5). Health related quality of life had a significant and clinically meaningful improvement from baseline to all follow-up for both groups across all measures. While the difference between treatment groups was not clinically meaningful, patients in the Endo arm had statistically significant greater improvements on some measures (e.g. Vascular Quality of Life [VascuQoL]) but not others (e.g. European Quality of Life 5D [EQ-5D]) (6).

Given differences in clinical outcomes and heath related quality of life, the current analysis evaluated the cost-effectiveness of Endo versus Bypass for patients with CLTI using data from the BEST-CLI trial.

## METHODS

### Model Overview

We developed an individual-level continuous-time Markov model using the Hesim package in R, a validated modular and computationally efficient R package for health economic simulation modeling and decision analysis (7–9).The model (**Figure 1**) consisted of five health states representing the occurrence of the pre-specified and adjudicated clinical events from the BEST-CLI trial. These states included: (1) “post re-intervention,” to indicate major index limb reintervention such as a new bypass graft or jump/interposition graft revision), (2) “post amputation,” which represented above ankle amputation of the index limb, (3) “post non-fatal myocardial infarction (MI) or stroke,” (4) “death” and (5) “no event”. The cycle length for the model was one year, during which patients could transition between the health states.

**Figure 1.**
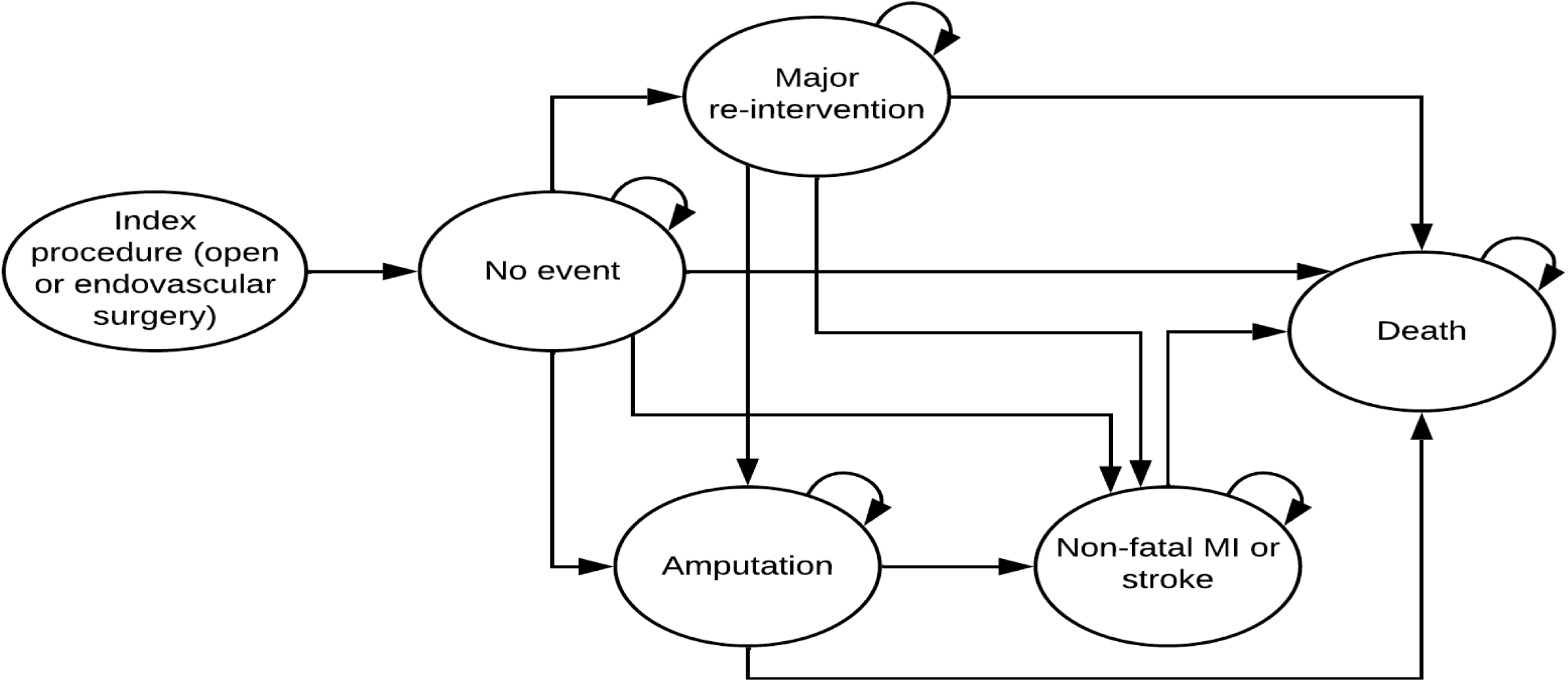
Schematic representation of the cost-effectiveness model.

### Model parameters

The model parameters (**Table 1**), including transition probabilities, resource utilization estimates and health-state utility values, were derived directly from “Cohort 1” of the BEST-CLI trial. As defined above, Cohort 1 consisted of individuals with an adequate single-segment great saphenous vein (SSGSV) and included 1434 of the 1830 patients enrolled in the trial.

**Table 1.** Input parameters for the model evaluating the cost-effectiveness of endovascular versus open surgery for chronic limb-threatening ischemia.

| Parameter | Base case estimate |  | Probability distribution | Source |
| --- | --- | --- | --- | --- |
| **Demographics** |  |  | Normal (for continuous variables) and binomial (for binary variables) distributions | RCT data |
| Age, mean years (SD) | 67 (10) |  |  |  |
| Female, % | 28% |  |  |  |
| BMI, mean (SD) | 28.5 (6.2) |  |  |  |
| **Comorbidities (prevalence)** |  |  |  |  |
| Coronary artery disease | 0.42 (0.49) |  |  |  |
| Diabetes | 0.73 (0.45) |  |  |  |
| Chronic heart failure | 0.06 (0.23) |  |  |  |
| Chronic kidney disease | 0.28 (0.45) |  |  |  |
| COPD | 0.15 (0.36) |  |  |  |
| Hypertension | 0.75 (0.44) |  |  |  |
| Stroke | 0.13 (0.34) |  |  |  |
| **Baseline medication use (prevalence)** |  |  |  |  |
| Direct oral anticoagulant | 0.04 (0.19) |  |  |  |
| Use of warfarin | 0.07 (0.25) |  |  |  |
| **Transition probabilities (annual)** | **Open surgery** | **Endovascular surgery** | Log-normal | RCT data |
|  | Mean (Standard error) | Mean (Standard error) |  |  |
| No event to re-intervention | 0.030 (0.008) | 0.079 (0.031) |  |  |
| No event to amputation | 0.024 (0.007) | 0.030 (0.012) |  |  |
| No event to MACE | 0.045 (0.012) | 0.053 (0.021) |  |  |
| No event to death | 0.051 (0.012) | 0.050 (0.018) |  |  |
| Re-intervention to amputation | 0.060 (0.017) | 0.076 (0.031) |  |  |
| Re-intervention to MACE | 0.045 (0.012) | 0.053 (0.021) |  |  |
| Re-intervention to death | 0.102 (0.024) | 0.101 (0.036) |  |  |
| Amputation to MACE | 0.045 (0.012) | 0.053 (0.021) |  |  |
| Amputation to death | 0.116 (0.027) | 0.115 (0.041) |  |  |
| MACE to death | 0.153 (0.036) | 0.151 (0.055) |  |  |
| **Health state utilities** |  |  | Beta | RCT data |
| No event (post index surgery) | 0.656 (0.196) | 0.683 (0.185) |  |  |
| Post re-intervention | 0.615 (0.250) | 0.672 (0.207) |  |  |
| Post amputation | 0.470 (0.274) | 0.416 (0.271) |  |  |
| Post non-fatal MI or stroke | 0.539 (0.261) | 0.602 (0.244) |  |  |
| **Index surgery costs** | \$14,273 | \$16,084 | Gamma | RCT data,<br>publicly available<br>data and published<br>literature (10–14) |
| **Maintenance costs (mean annual)** |  |  |  |  |
| No event (post index surgery) | \$44,558 (8,912) | \$44,558 (8,912) | | |
| Post re-intervention | \$66,668 (13,334) | \$66,668 (13,334) | | |
| Post amputation | \$176,315<br>(35,263) | \$176,315<br>(35,263) | | |
| Post non-fatal MI or stroke | \$74,514 (14,903) | \$74,514 (14,903) | | |

RCT: randomized controlled trial; BMI: body mass index; COPD: chronic obstructive pulmonary disease; MACE: major adverse cardiovascular event.

#### Transition probabilities

We used Cox proportional hazards models to estimate the hazards of major re-intervention, above-ankle amputation, non-fatal stroke or MI, and death from the BEST-CLI trial data as a function of baseline covariates including treatment arm (i.e. Bypass v. Endo), age, sex, body mass index (BMI), hypertension, diabetes mellitus, prior stroke, coronary artery disease (CAD), congestive heart failure (CHF), chronic kidney disease (CKD), chronic obstructive pulmonary disease (COPD), use of a direct oral anticoagulant and use of warfarin. These covariates were selected using Least Absolute Shrinkage and Selection Operator (LASSO) regressions along with clinical judgment. We validated these prediction models using the rms package in R. The exponent of the hazard ratio represents the time to event for each outcome for individual patients. We display the average value of these probabilities in **Table 1**.

#### Direct medical costs

We used BEST-CLI data to estimate healthcare resource utilization. Our analysis was conducted from the 2024 US payer perspective. For each health state in the model in a given treatment arm, we estimated the number of annualized hospitalization days (10), intensive care unit (ICU) days (11), emergency room (ER) visits (12), dialysis (13), tracheostomy, gastrointestinal (GI) endoscopy, cardiac catheterizations, computed tomography (CT) scans, magnetic resonance imaging (MRI) scans, ultrasounds, echocardiograms, cardiac stress tests, physical therapy sessions, outpatient visits, outpatient rehabilitation hours, and inpatient rehabilitation hours (14). We then multiplied estimates of these resources by their respective unit costs, derived from publicly available data sources (10–14), to estimate the average direct medical costs associated with each health state in each treatment arm. The costs of index procedures were based an analysis of Centers for Medicare and Medicaid Services (CMS) claims data representing costs for the procedure and the next 30 days. The resulting costs estimates are reported in **Table 1**.

#### Health state utility values

We used EQ-5D (15,16) scores, a self-reported general health related quality of life index, collected during BEST-CLI to calculate the mean and standard error (SE) of utility values associated with each of five model health states in each treatment arm. The utility estimates are reported in **Table 1**.

### Base case analysis

For the base case, we conducted our analyses over time horizons of five and ten years. All future costs and quality-adjusted life year (QALY) gains were discounted at 3% consistently with recommendations of the US Second Panel on Cost-effectiveness in Health and Medicine (17). We calculated the cost-effectiveness of Endo vs. Bypass surgery in terms of both their incremental costs per life year gained and incremental costs per quality-adjusted life year (QALY) gained.

We also estimated the incremental net monetary benefit (INMB) of Endo vs. Bypass, which was calculated by multiplying the incremental QALYs gained by a willingness-to-pay value of $100,000/QALY and then subtracting from this product the incremental costs. Additionally, we calculated the incremental costs to avoid the individual health states in the model, i.e., major intervention, above-ankle amputation and non-fatal MI or stroke.

### Sensitivity analyses

We performed extensive sensitivity analyses based on assumed probability distributions for the model input parameters (**Table 1**). We conducted one-way sensitivity analyses on each of the input parameters, including patient characteristics (i.e., age, sex, body mass index [BMI], and presence of major baseline comorbidities) transition probabilities, health-state utility values, and medical costs. We summarized these results using a tornado diagram.

We conducted probabilistic analyses using a Monte Carlo simulation with 10,000 iterations. Model input parameters were randomly drawn from their probability distributions in each iteration, and the outcomes were estimated for each set of input parameters. We presented the results of the probabilistic analyses using 95% credibility intervals (95% CrI) covering 95% of the random simulations. In addition, we plotted the results of the probabilistic analyses in a cost-effectiveness plane and a cost-effectiveness acceptability curve.

## RESULTS

### Model calibration

Our simulation model closely replicated the observed results from the BEST-CLI trial over the five-year follow-up (**Online Supplement, Figures S1**). Similarly, the observed relative risks of the four clinical outcomes in the trial fell within the 95% credible intervals of the simulation model’s predicted relative risks (**Online Supplement, Figure S2**).

### Base case results

Results for our base case analyses are shown in **Table 2**. For a 5-year time horizon, patients treated with Bypass were estimated to have direct medical costs of $227,341 (95% CrI: $173,075, $291,443) and a mean survival of 3.91 years (95% CrI: 3.78, 4.03). Patients treated with Endo had direct medical costs of $243,614 (95% CrI: $190,112, $305,605) and a mean survival of 3.88 years (95% CrI: 3.68, 4.06). As a consequence, Endo was dominated by Bypass surgery (i.e. Bypass was more effective and less costly than Endo) in terms of the incremental costs per life year gained.

**Table 2.** Main results for the cost-effectiveness of endovascular vs open surgery for chronic limb-threatening ischemia.

| | Open surgery | Endovascular surgery | Difference (Endovascular minus Open) | Incremental cost per life year gained (\$/LY) | Incremental cost per quality-adjusted-life year gained (\$/QALY) | Incremental net monetary benefit, \$ (95% CI) |
| --- | --- | --- | --- | --- | --- | --- |
| 5-year time horizon |  |  |  |  |  |  |
| Cost, mean \$ (95% CI) | 227,341 (173,075 to 291,443) | 243,614 (190,112 to 305,605) | 16,274 (-2,564 to 38,825) | Dominated | 263,973 | -10,109 (-168,908 to -157,433) |
| Survival, mean years (95% CI) | 3.91 (3.78, 4.03) | 3.88 (3.68, 4.06) | -0.030 (-0.184, 0.115) |  |  |  |
| Quality-adjusted life years, mean (95% CI) | 2.48 (1.11, 3.49) | 2.54 (1.39, 3.40) | 0.062 (-1.532, 1.731) |  |  |  |
| 10-year time horizon |  |  |  |  |  |  |
| Cost, mean \$ (95% CI) | 383,769 (298,663 to 481,969) | 414,316 (324,747 to 515,605) | 30,546 (-9,217 to 77,544) | Dominated | 1,104,753 | -27,781 (-255,103 to 212,390) |
| Survival, mean years (95% CI) | 6.18 (5.88, 6.47) | 6.05 (5.58, 6.49) | -0.127 (-0.468, 0.196) |  |  |  |
| Quality-adjusted life years, mean (95% CI) | 3.85 (1.89, 5.38) | 3.87 (2.30, 5.19) | 0.028 (-2.232, 2.410) |  |  |  |

QALYs: quality-adjusted life years; NMB: net monetary benefit (calculated from multiplying the willingness-to-pay value of \$100,000/QALY by incremental QALYs gained minus incremental costs); ICER: incremental cost-effectiveness ratio.

The mean QALYs per person were 2.48 (95% CrI: 1.11, 3.49) and 2.54 (95% CrI: 1.39, 3.40) for Bypass, and Endo, respectively. This resulted in an incremental costs per QALY gained of $263,973/QALY and an INMB of –$10,109 (95% CrI: –$168,908, $157,433) for Endo vs. Bypass surgery.

Results for over a 10-year time horizon were similar to the 5-year results. Endo was dominated by Bypass with respect to incremental costs per life year gained and had an incremental costs per QALY gained of $1,104,753/QALY and an INMB of –$27,781 (95% CrI: –$255,103, $212,390).

Results in terms of major events are presented in **Online Supplement, Table S1**. Over a 5-year time horizon, Bypass reduced spending by $117,466 (95% CrI: –$368,099, $20,702) for every major reintervention avoided, reduced spending by $463,257 (95% CrI: –$3,528,089, $1,367,655) for every amputation avoided, and reduced spending by $614,346 (95% CrI: –8,966,490, $5,186,083) for every MI or stroke avoided. Results over a 10-year time horizon were similar.

### Sensitivity analyses

Figure 2 shows the results of the one-way sensitivity analysis over a five-year time horizon. In all one-way sensitivity analyses, INMB values were negative, meaning that Bypass remained more cost-effective than Endo at a willingness to pay threshold of $100,000/QALY. The model was most sensitive to the cost of re-intervention, baseline age, and health-state utility value of the post re-intervention state. Changing the cost of re-intervention by 20% from its base case value changed the INMB from a low of – $13,946 to a high of –$6,272. Changing the mean baseline age of patients by a standard deviation from its base case value changed the INMB from a low of –$14,822 to a high of –$6,300. Changing the health state utility value of the post re-intervention state by 20% changed the INMB from a low of –$15,434 to a high of –$5,851.

**Figure 2.**
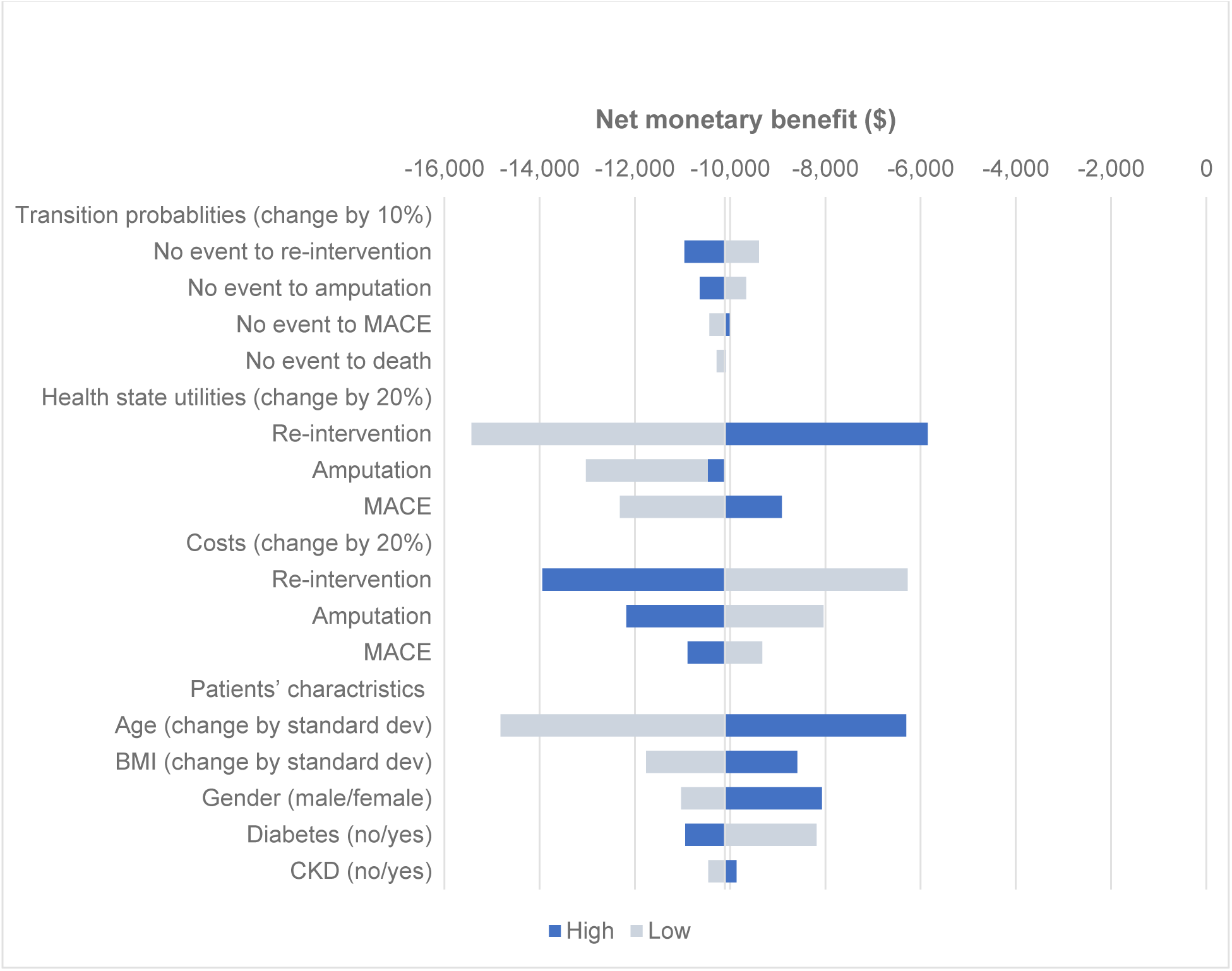
One-way sensitivity analyses for the cost-effectiveness of endovascular vs open surgery for chronic limb-threatening ischemia.

Figure 3 shows the results of the probabilistic analysis (**Panels A-C**). At a willingness-to-pay threshold of $100,000/QALY, Bypass was more cost-effective than Endo in 55% of the Monte Carlo simulations. The probability of cost-effectiveness of Bypass reduced to 52% at a willingness-to-pay threshold of $200,000/QALY. For willingness to pay values larger than $350,000/QALY, Endo was slightly more cost-effective than Bypass (51% vs. 49%).

**Figure 3.**
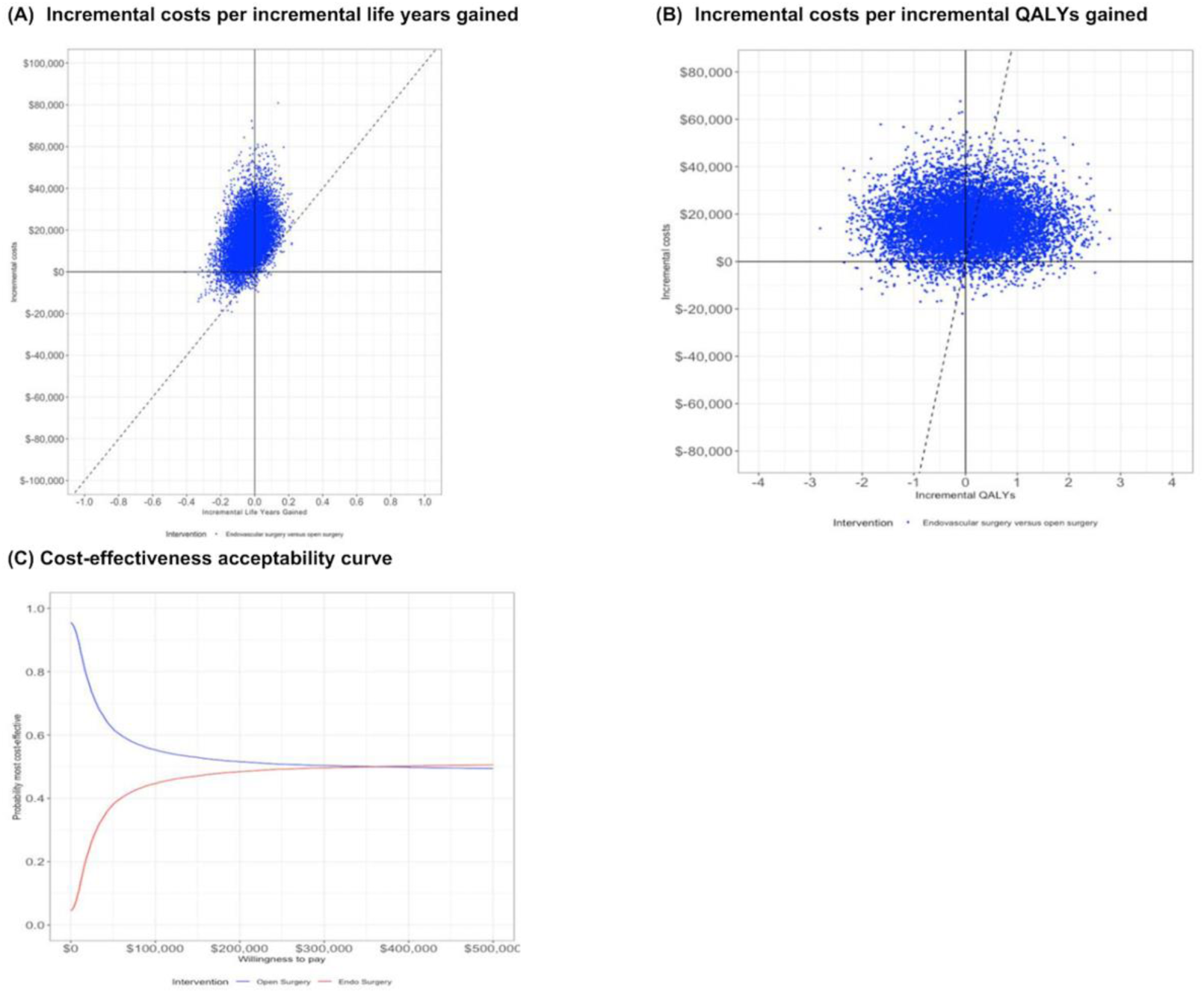
Probabilistic analyses results for the cost-utility analysis of endovascular vs open surgery for chronic limb-threatening ischemia. Willingness-to-pay at $100,000/QALY.

## DISCUSSION

We assessed the cost-effectiveness of Endo compared with Bypass surgery for patients with CLTI by conducting an individual level simulation model using data from the BEST-CLI trial. Our analysis has three primary findings.

First, when considering the incremental costs of each additional year of survival, Bypass was associated with lower costs and better survival over both a 5 and 10-year time horizon in our base case results, meaning that Bypass was economically “dominant” compared with Endo. Consistent with this, every major clinical event avoided by Bypass was associated with large cost savings.

Second, when health-related quality of life was incorporated into the measures of effectiveness in our base case results, Endo resulted in slightly higher quality-adjusted life years (QALYs) than Bypass but with higher costs. The resultant incremental cost per additional QALY gained were substantially above the conventional willingness-to-pay threshold of $100,000/QALY at both 5 and 10 years. This means that Bypass would remain the cost-effective approach at this threshold.

Third, we found substantial uncertainty around the base case estimates. When we conducted the probabilistic Monte Carlo simulation, the 95% credible interval for the INMB ranged from very negative (i.e. Bypass is the clearly preferred approach) to very positive (i.e. Endo is likely cost-effective). Consistent with this, at a willingness to pay threshold of $100,000/QALY, there is only a 55% chance that Bypass is more cost-effective than Endo. Given the substantial uncertainty around model parameters, there is likely substantial “value of information” that could be gained from better characterizing the relative effects and costs of these two revascularization approaches for CLTI (18).

Only two prior randomized control trials have evaluated the cost-effectiveness of open and endovascular revascularization approaches for patients with CLTI(19). In contrast to our results, a cost-utility analysis of the Bypass versus Angioplasty in Severe Ischaemia of the Leg (BASIL) trial (20) found that Bypass was associated with slightly higher spending and greater health related adjusted survival, with a resultant incremental cost per QALY gained of $184,492 per QALY(21). A cost-utility analysis of the more recently completed BASIL 2 trial found that Bypass was associated with substantially higher costs and slightly higher QALYs than Endo with a resultant incremental cost per QALY gained for Bypass compared with Endo of $730,000 per QALY gained (22). Other results from these models were more similar to our findings: the cost effectiveness analysis of BASIL found substantial uncertainty in the results of their base case analyses, with 35% of their probabilistic analyses finding Endo to be more effective than Bypass, and the cost effectiveness analysis of BASIL 2 found substantial cost savings per amputation avoided.

There are numerous challenges comparing the results of these prior trials with those from BEST-CLI. Both of these trials were conducted in the UK. BASIL was completed more than 20 years ago, during which time endovascular treatment was mostly done with balloon angioplasty, quality of life data was missing for about one-third of patients and the analysis only considered in-hospital costs. BASIL 2 trial participants were older than those in BEST and all had infra-popliteal revascularization; in contrast, 42% of BEST-CLI patients had femoro-popliteal procedures. Both of the BASIL cost-effectiveness analyses considered shorter time horizons than we evaluated and were conducted by taking cohort-level data observed during these trials. In contrast, we built our Markov model at the individual level, which enables prediction of outcomes for different patient subgroups. We looked at a wide set of covariates that predict the clinical outcomes, including re-intervention, amputation, and non-fatal MI or stroke, and chose covariates both based on their clinical relevance and statistical performance.

It is important to acknowledge some of the limitations of our approach. We focused our analysis on the components of the primary efficacy endpoint (major reintervention, major amputation and death) and the adjudicated safety endpoints of MI and stroke. Patients with CLTI frequently undergo minor reinterventions (e.g. angioplasty or stenting), receive wound care and debridements and have minor amputations. Post-hoc analyses from BEST-CLI have demonstrated that patients randomized to Bypass had fewer total vascular reinterventions per patient (major or minor) (23), fewer total amputation events (major or minor), and reduced incidence of recurrent CLTI (24,25). Inclusion of these outcomes and their associated costs is likely to have made Bypass appear even more favorable and may have resulted in it being the cost-effective strategy in even a greater proportion of probabilistic simulation scenarios than we found.

In summary, our analysis demonstrates that Bypass delivers greater survival and a lower rate of major adverse limb events at a lower cost than Endo. Further, while Endo results in greater health related quality of life, these incremental benefits represent low value for money based on conventional willingness to pay thresholds. Perhaps most importantly, while these results taken together favor Bypass, we found substantial uncertainty in our estimates, justifying future research to identify subgroups for whom each of these approaches may definitively be cost-effective.

## Data Availability

The individual participant data from the BEST-CLI randomized clinical trial used in this study are not publicly available because of patient privacy restrictions and trial data use agreements. Requests for access to the BEST-CLI data should be directed to the trial investigators. The cost-effectiveness model developed in this study and the aggregated model inputs, assumptions, and outputs are available from the corresponding author upon request.

## Acknowledgements

**Funding support:** BEST-CLI was supported by the National Heart, Lung, and Blood Institute of the National Institutes of Health under Award Numbers U01HL107407, U01HL107352, and U01HL115662. The following entities also provided funding to the BEST-CLI trial during the follow-up period (2019–2021). These include **Physician Societies:** Vascular InterVentional Advances (VIVA), Society for Vascular Surgery, New England Society for Vascular Surgery, Western Vascular Society, Eastern Vascular Society, Midwest Vascular Surgery Society, Southern Association of Vascular Surgeons, Canadian Society for Vascular Surgery, Society for Clinical Vascular Surgery, Society of Interventional Radiology, Vascular and Endovascular Surgery Society, Society for Vascular Medicine; and **Industry sources:** Janssen, Gore, Becton Dickinson and Company, Medtronic, Cook, Boston Scientific, Abbott, Cordis, Cardiovascular Systems Inc. As of September 1, 2022, ongoing BEST-CLI research is funded primarily by Grant Number NNF22SA0078610 from the Novo Nordisk Foundation.

## Originality of Content

All information in the manuscript is original.

## SUPPLEMENTAL INFORMATION

**Table S1.** Results of cost-effectiveness analysis presented as incremental costs per clinical event avoided.

| | Open surgery | Endovascular surgery | Difference (Endovascular minus Open) | Incremental costs per major re-intervention avoided, \$ (95% CI) | Incremental costs per amputation avoided, \$ (95% CI) | Incremental costs per non-fatal MI or stroke avoided, \$ (95% CI) |
| --- | --- | --- | --- | --- | --- | --- |
| 5-year time horizon |  |  |  |  |  |  |
| Cost, mean \$ (95% CI) | 227,341 (173,075 to 291,443) | 243,614 (190,112 to 305,605) | 16,274 (-2,564 to 38,825) | -117,466 (-368,099 to 20,702) | -463,257 (-3,528,089 to 1,367,655) | -614,346 (-8,966,490 to 5,186,083) |
| Rate of major re-intervention | 0.109 (0.075, 0.149) | 0.247 (0.141, 0.387) | 0.139 (0.054, 0.248) |  |  |  |
| Rate of above ankle amputation | 0.103 (0.070, 0.139) | 0.138 (0.080, 0.215) | 0.035 (-0.008, 0.090) |  |  |  |
| Rate of non-fatal MI and stroke | 0.180 (0.135, 0.231) | 0.206 (0.134, 0.301) | 0.026 (-0.025, 0.089) |  |  |  |
| 10-year time horizon |  |  |  |  |  |  |
| Cost, mean \$ (95% CI) | 383,769 (298,663 to 481,969) | 414,316 (324,747 to 515,605) | 383,769 (298,663 to 481,969) | -184,600 (-582,910 to 66,030) | -591,430 (-3,179,380 to 994,000) | -947,850 (-13,587,270 to 8,270,790) |
| Rate of major re-intervention | 0.159 (0.114, 0.211) | 0.324 (0.192, 0.479) | 0.165 (0.066, 0.279) |  |  |  |
| Rate of above ankle amputation | 0.158 (0.115, 0.207) | 0.210 (0.131, 0.308) | 0.052 (-0.006, 0.120) |  |  |  |
| Rate of non-fatal MI and stroke | 0.273 (0.215, 0.335) | 0.305 (0.214, 0.413) | 0.032 (-0.030, 0.101) |  |  |  |

**Figure S1.**
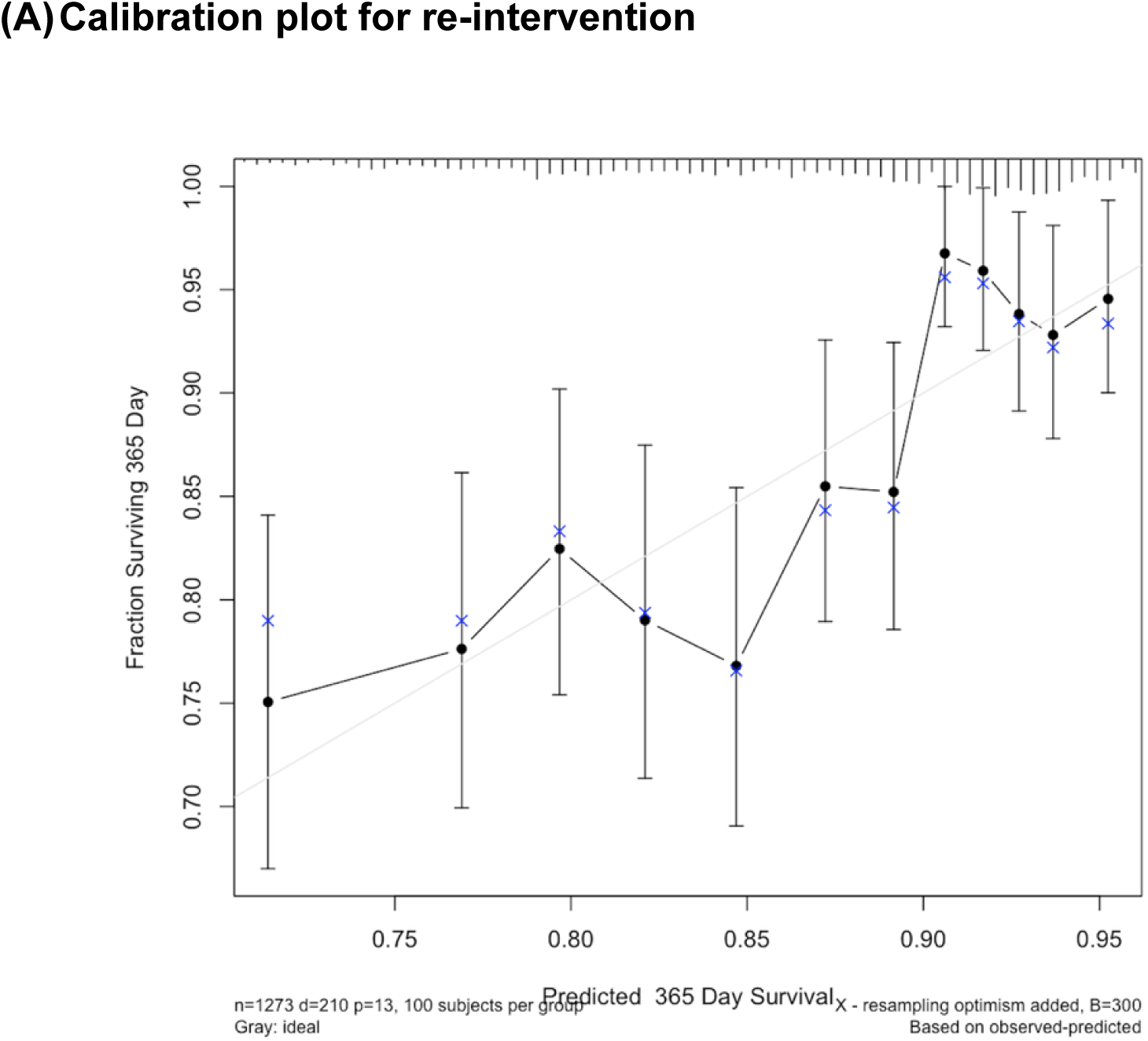
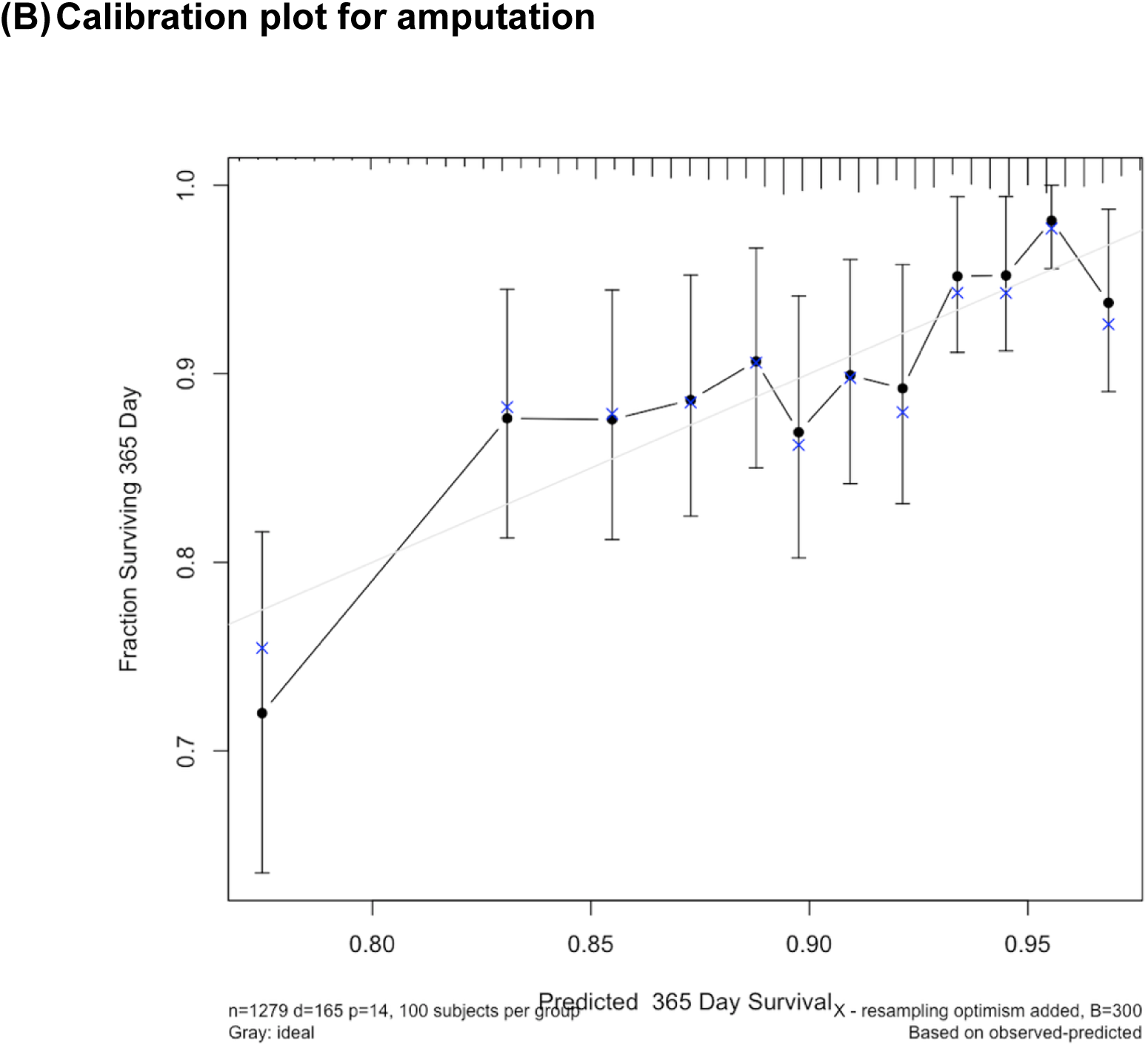
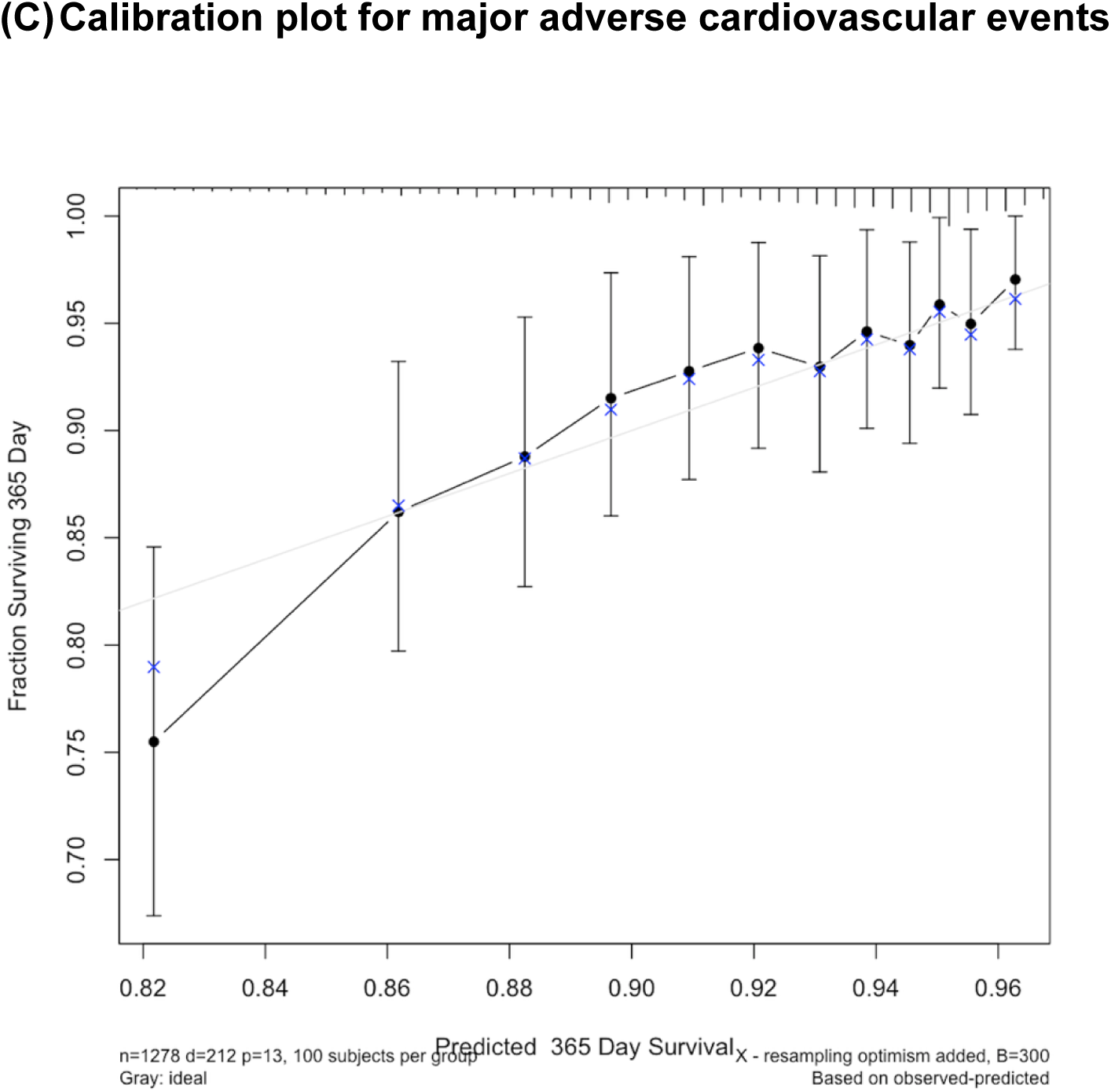
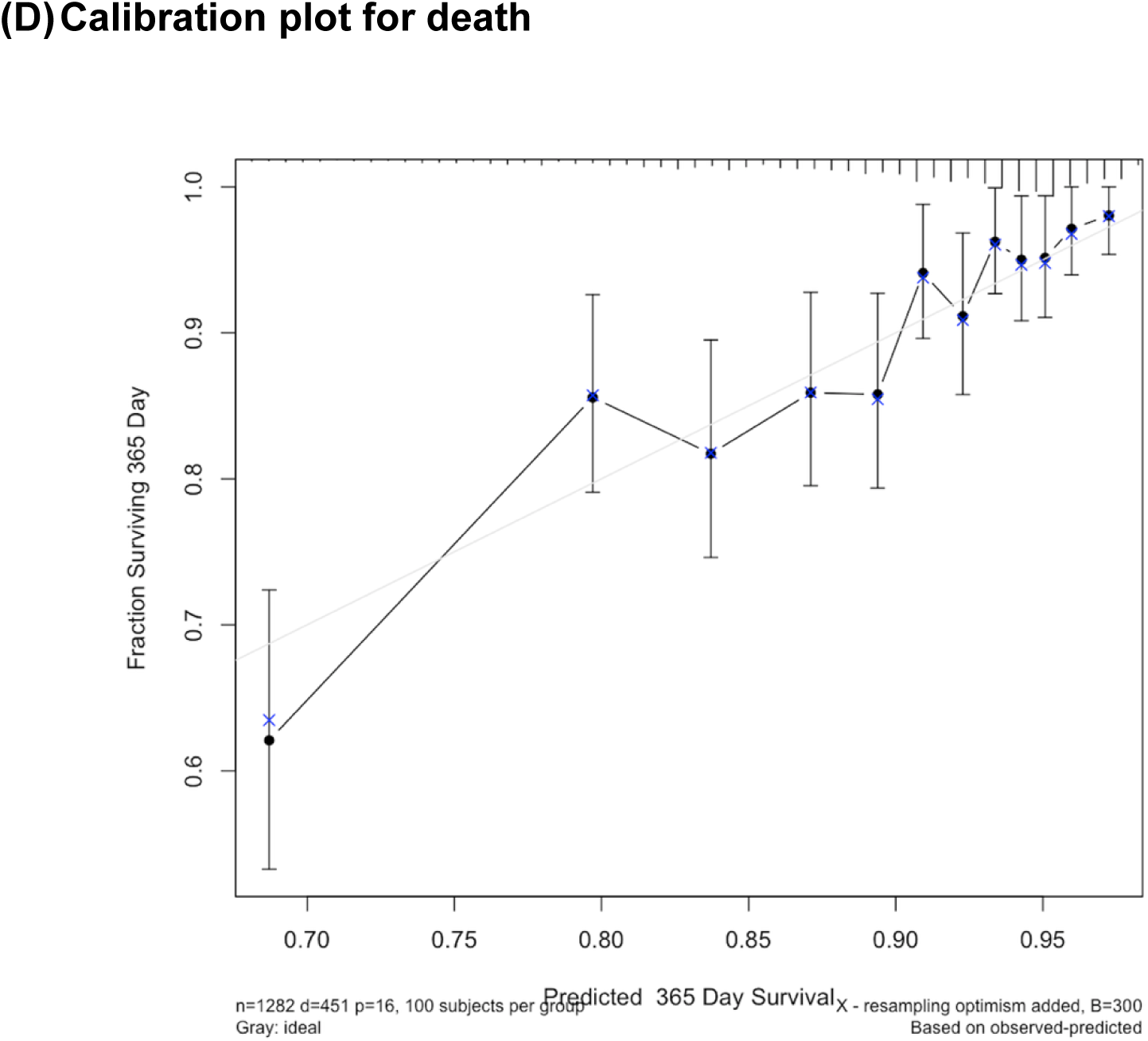
Calibration plots for trial observed versus regression predicted probabilities of the clinical outcomes (re-intervention, amputation, MACE, and death) within 365 days.

**Figure S2.**
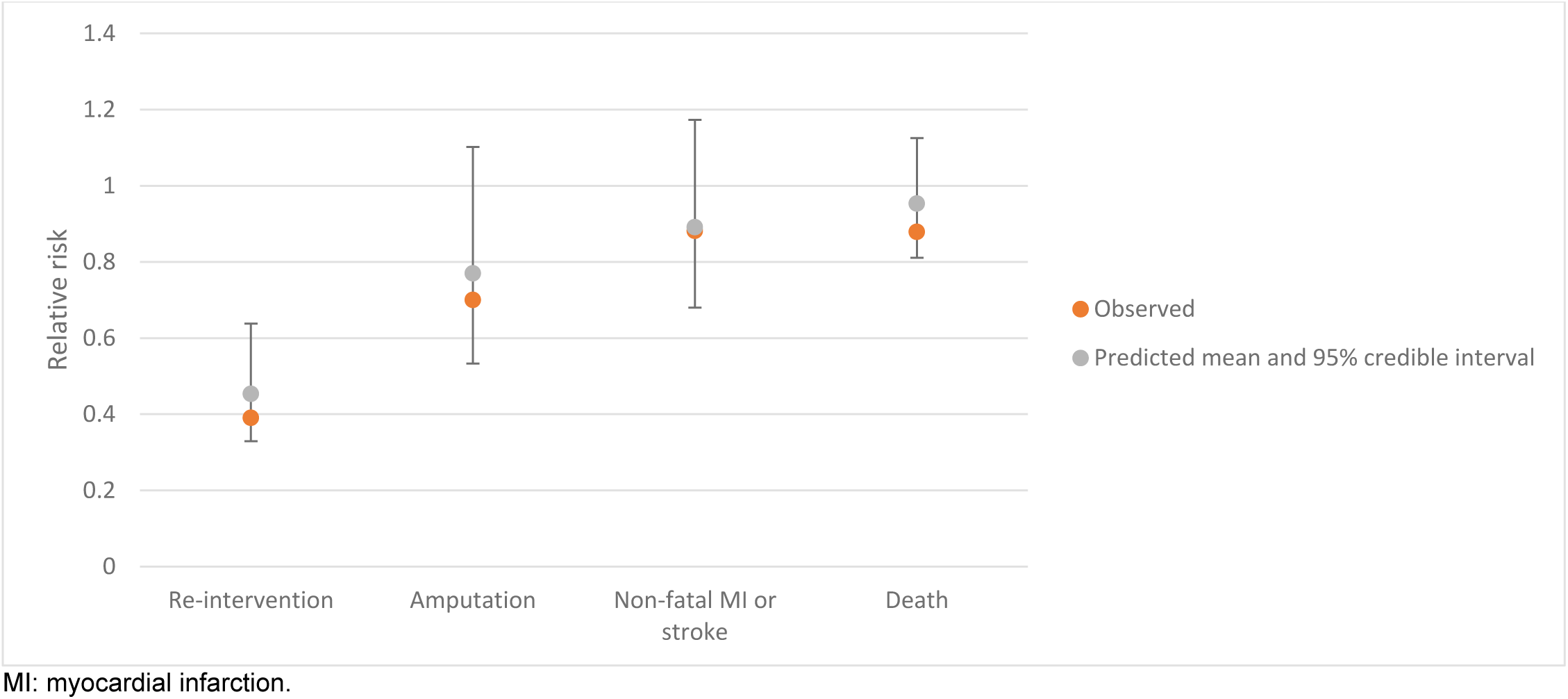
Trial observed versus model predicted relative risks of the clinical outcomes (re-intervention, amputation, non-fatal MI or stroke, and death) for Bypass versus Endo.

